# Analysis of 50,000 exome-sequenced UK Biobank subjects fails to identify genes influencing probability of psychiatric referral

**DOI:** 10.1101/2020.07.16.20155267

**Authors:** David Curtis

## Abstract

**Background:** Depression is moderately heritable but there is no common genetic variant which has a major effect on susceptibility. It is possible that some very rare variants could have substantial effect sizes and these could be identified from exome sequence data.

**Methods:** Data from 50,000 exome-sequenced UK Biobank participants was analysed. Subjects were treated as cases if they had reported having seen a psychiatrist for “nerves, anxiety, tension or depression”. Gene-wise weighted burden analysis was performed to see if there were any genes or sets of genes for which there was an excess of rare, functional variants in cases.

**Results:** There were 5,872 cases and 43,862 controls. There were 22,028 informative genes but none produced a statistically significant result after correction for multiple testing. Of the 25 genes individually significant at p<0.001 none appeared to be a biologically plausible candidate. No set of genes achieved statistical significance after correction for multiple testing and those with the lowest p values again did not appear to be biologically plausible candidates.

**Limitations:** The phenotype is based on self-report and the cases are likely to somewhat heterogeneous. The number of cases is on the low side for a study of exome sequence data.

**Conclusions:** The results conform exactly with the expectation under the null hypothesis. It seems unlikely that depression genetics research will produce findings that might have a substantial clinical impact until far larger samples become available.

## Introduction

Depression is moderately heritable and genome wide association studies have demonstrated that no common genetic variant has a major effect on susceptibility (Howard et al., 2019). It remains possible that there could be very rare variants with major effect size and these might be identified from analysis of exome sequence data.

The UK Biobank is a resource of 500,000 British volunteers who have undergone extensive phenotyping and who have donated biological samples, including DNA sample (http://www.ukbiobank.ac.uk/about-biobank-uk/). They are intended to be broadly representative of the population, though targeted to have a higher than average age, and because depression is a relatively common diagnosis it is expected that many subjects with a lifetime history of depression will be included. As discussed recently, when selecting an appropriate definition for genetic studies there is a trade-off between focussing on more narrowly defined severe cases and broadening the criteria to include more subjects at the cost of the sample become more heterogeneous (Ormel et al., 2019). This is especially the case for biobank-based studies rather than case-control studies. Another consideration is that there are a number of different depression-related fields in the UK Biobank data but not all of them have entries for all subjects. For example, the item “Number of depression episodes” is available for only 90,000 subjects. Some information is comprehensively recorded, such as hospital admissions and prescribed medication. On the other hand, very few patients with depression are admitted to hospital whereas large numbers of people might be prescribed antidepressants at one time or another for indications which might be quite mild and non-specific. At initial assessment, all participants fill out a touchscreen questionnaire and one item is “Have you ever seen a psychiatrist for nerves, anxiety, tension or depression?” to which 497,000 answered either Yes or No. In the UK most cases of depression and anxiety are treated by general practitioners so patients who had been referred to a psychiatrist would be expected to be significantly unwell with some kind of mood disorder.

Exome sequence data has been released for 50,000 subjects so a gene-based weighted burden analysis was carried out to determine if there were any genes for which an increased burden of rare and/or functionally significant variants was associated with increased probability of having been referred for psychiatric treatment.

## Methods

The same approach was followed as has been previously described (Curtis, 2020a).The UK Biobank dataset was downloaded along with the variant call files for 49,953 subjects who had undergone exome-sequencing and genotyped using the GRCh38 assembly with coverage 20X at 94.6% of sites on average (Hout et al., 2019). UK Biobank had obtained ethics approval from the Research Ethics Committee (REC; approval number: 11/NW/0382) and informed consent from all participants. All variants were annotated using VEP, PolyPhen and SIFT (Adzhubei et al., 2013; Kumar et al., 2009; McLaren et al., 2016). To obtain population principal components reflecting ancestry, version 1.90beta of *plink* (https://www.cog-genomics.org/plink2) was run with the options *--maf 0*.*1 --pca header tabs --make-rel* (Chang et al., 2015; Purcell et al., 2007).

The phenotype was determined according to how participants had responded in their initial assessment to the touchscreen question: “Have you ever seen a psychiatrist for nerves, anxiety, tension or depression?” Those answering “Yes” were taken to be cases and those answering “No” controls.

SCOREASSOC was used to carry out a weighted burden analysis to test whether, in each gene, sequence variants which were rarer and/or predicted to have more severe functional effects occurred more commonly in cases than controls. Attention was restricted to rare variants with minor allele frequency (MAF) <= 0.01. As previously described, variants were weighted by MAF so that variants with MAF=0.01 were given a weight of 1 while very rare variants with MAF close to zero were given a weight of 10 (Curtis, 2020b). Variants were also weighted according to their functional annotation using the default weights provided with the GENEVARASSOC program, which was used to generate input files for weighted burden analysis by SCOREASSOC (Curtis, 2016, 2012). As previously described, the weighting scheme ranges from 1 for intergenic variants to 20 for gene-disruptive variants (Curtis, 2020b). Additionally, 10 was added to the weight if the PolyPhen annotation was possibly or probably damaging and also if the SIFT annotation was deleterious, meaning that a non-synonymous variant annotated as both damaging and deleterious would be assigned an overall weight of 30. The weight due to MAF and the weight due to functional annotation were then multiplied together to provide an overall weight for each variant. Variants were excluded if there were more than 10% of genotypes missing in the controls or if the heterozygote count was smaller than both homozygote counts in the controls. If a subject was not genotyped for a variant then they were assigned the subject-wise average score for that variant. For each subject a gene-wise weighted burden score was derived as the sum of the variant-wise weights, each multiplied by the number of alleles of the variant which the given subject possessed. For variants on the X chromosome, hemizygous males were treated as homozygotes.

For each gene, a ridge regression analysis was carried out with lamda=1 to test whether the gene-wise variant burden score was associated with the hyperlipidaemia phenotype. To do this, SCOREASSOC first calculates the likelihood for the phenotypes as predicted by the first 20 population principal components and then calculates the likelihood using a model which additionally incorporates the gene-wise burden scores. It then carries out a likelihood ratio test assuming that twice the natural log of the likelihood ratio follows a chi-squared distribution with one degree of freedom to produce a p value. The statistical significance is summarised as a signed log p value (SLP) which is the log base 10 of the p value given a positive sign if the score is higher in cases and negative if it is higher in controls. We have shown that incorporating population principal components in this way satisfactorily controls for test statistic inflation when applied to this heterogeneous dataset (Curtis, 2020b).

Gene set analyses were carried out using the 1454 “all GO gene sets, gene symbols” pathways as listed in the file *c5*.*all*.*v5*.*0*.*symbols*.*gmt* downloaded from the Molecular Signatures Database at http://www.broadinstitute.org/gsea/msigdb/collections.jsp (Subramanian et al., 2005). For each set of genes, the natural logs of the gene-wise p values were summed according to Fisher’s method to produce a chi-squared statistic with degrees of freedom equal to twice the number of genes in the set. The p value associated with this chi-squared statistic was expressed as a minus log10 p (MLP) as a test of association of the set with the mental illness phenotype.

## Results

There were 5,872 cases who reported that they had seen a psychiatrist for a mental health problem and 43,862 controls. There were 22,028 genes for which there were qualifying variants and the QQ plot for the SLPs obtained for each gene is shown in Figure 1. This shows that the test is well-behaved and that the SLPs conform exactly to what one would expect under the null hypothesis that there are no genes for which an increased burden of rare, functional variants affects the risk of mental illness.

**Figure 1.**
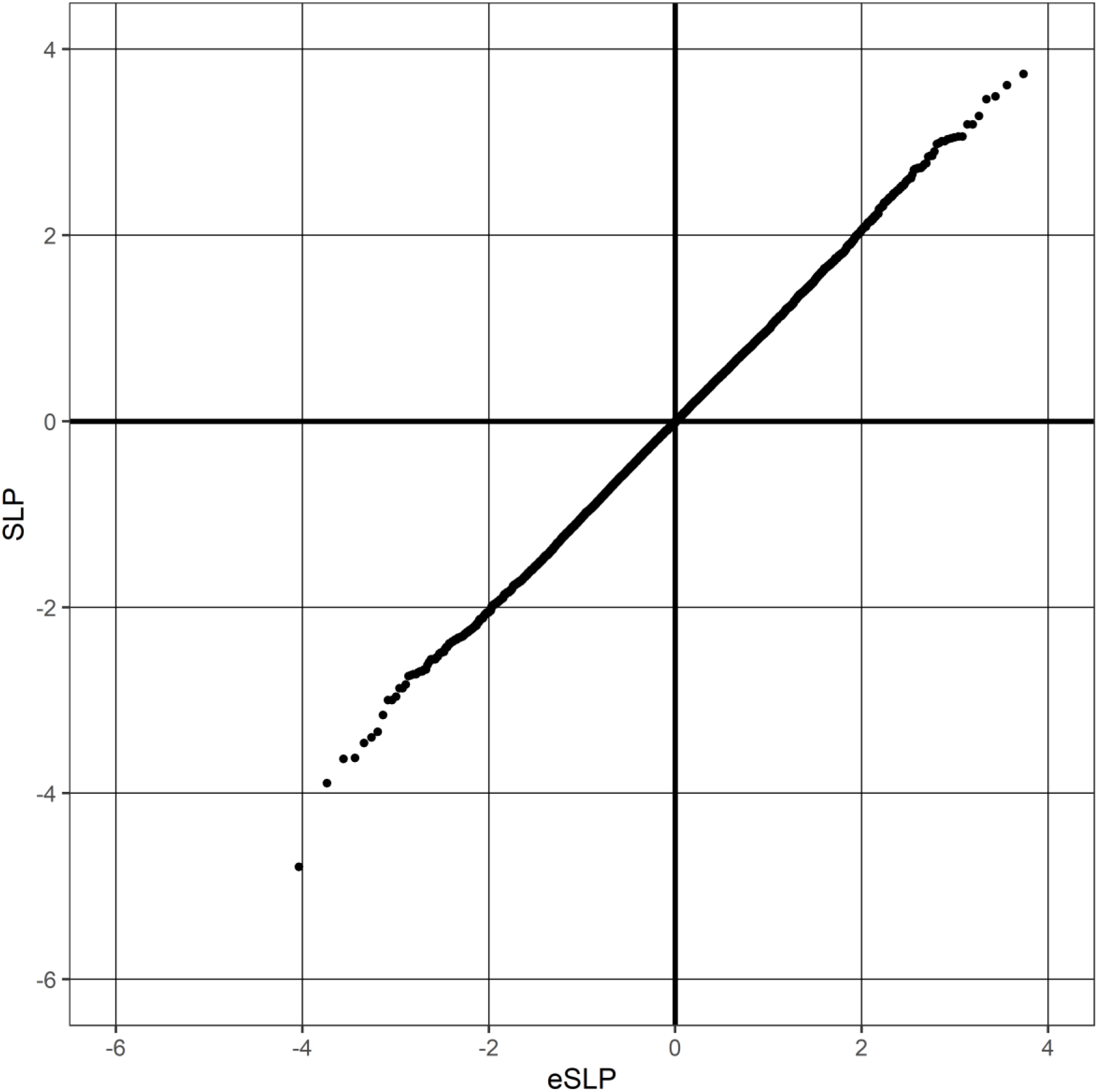
QQ plot of SLPs obtained for weighted burden analysis of 22,028 genes for association with psychiatric treatment showing observed against expected SLP for each gene.

Table 1 shows the results for all genes with an absolute value of SLP >= 3 (equivalent to p<=0.001). By chance, from 22,028 genes one would expect 11 to have SLP greater than 3 and 11 to have SLP less than -3, whereas the actual numbers are 15 and 10. Applying a Bonferroni correction to test for genome-wide statistical significance would yield a threshold of log10(22,028/0.05)=5.6 for the absolute value of the SLP and no gene achieves this. The genes listed in the table are involved in a wide variety of different functions and none appears to be a plausible biological candidate for affecting susceptibility to depression or anxiety.

**Table 1.** Genes with absolute value of SLP exceeding 3 or more (equivalent to p<0.001) for test of association of weighted burden score with psychiatric treatment.

| Gene symbol | SLP | Gene name |
| --- | --- | --- |
| *LCE2B* | 4.00 | Late Cornified Envelope 2B |
| *IRF3* | 3.73 | Interferon Regulatory Factor 3 |
| *COL11A1* | 3.61 | Collagen Type XI Alpha 1 Chain |
| *RPS3A* | 3.49 | Ribosomal Protein S3A |
| *FAM161B* | 3.46 | FAM161 Centrosomal Protein B |
| *BIN2* | 3.28 | Bridging Integrator 2 |
| *MRPL55* | 3.19 | Mitochondrial Ribosomal Protein L55 |
| *USP42* | 3.19 | Ubiquitin Specific Peptidase 42 |
| *TRAF1* | 3.06 | TNF Receptor Associated Factor 1 |
| *GCC2-AS1* | 3.06 | GCC2 Antisense RNA 1 |
| *PAGE5* | 3.05 | PAGE Family Member 5 |
| *CNOT9* | 3.04 | CCR4-NOT Transcription Complex Subunit 9 |
| *MC1R* | 3.03 | Melanocortin 1 Receptor |
| *KLK12* | 3.01 | Kallikrein Related Peptidase 12 |
| *PAF1* | 3.01 | PAF1 Homolog, Paf1/RNA Polymerase II Complex Component |
| *LOC105370163* | -3.00 | Uncharacterized LOC105370163 |
| *SPINK2* | -3.00 | Serine Peptidase Inhibitor Kazal Type 2 |
| *ABCD3* | -3.16 | ATP Binding Cassette Subfamily D Member 3 |
| *ZSWIM1* | -3.34 | Zinc Finger SWIM-Type Containing 1 |
| *TTLL4* | -3.40 | Tubulin Tyrosine Ligase Like 4 |
| *OR6B1* | -3.46 | Olfactory Receptor Family 6 Subfamily B Member 1 |
| *H2AFY2* | -3.62 | MacroH2A.2 Histone |
| *CREBZF* | -3.63 | CREB/ATF BZIP Transcription Factor |
| *MUC19* | -3.89 | Mucin 19, Oligomeric |
| *GADD45A* | -4.79 | Growth Arrest And DNA Damage Inducible Alpha |

There is no gene set which would be statistically significant after a Bonferroni correction for the number of sets tested. The most significant set was HYDROLASE ACTIVITY ACTING ON ACID ANHYDRIDES CATALYZING TRANSMEMBRANE MOVEMENT OF SUBSTANCES (MLP=3.05) which contains 35 genes coding for ATPases. Neither this set nor the others with relatively high MLPs appear to be biologically plausible candidates.

The SLPs for all genes and the MLPs for all gene sets are provided in supplementary tables S1 and S2.

## Discussion

The results are completely negative. No gene is formally statistically significant after correction for multiple testing and even those which are ranked highest and lowest do not include any which could be regarded as being biologically plausible candidates. The distribution of results is exactly as one would expect by chance. These results contrast with those obtained when the same methods were applied to similar sample sizes of other complex phenotypes, comprising schizophrenia, late onset Alzheimer’s disease, BMI and hyperlipidaemia (Curtis, 2020b, 2020a; Curtis et al., 2019, 2018). For these phenotypes it was possible to identify gene sets which were statistically significant and a small number of biologically plausible genes which were either genome-wide significant or which at least had uncorrected p values less than 0.001. In the present dataset there is not even a suggestion of such a signal.

## Limitations

The method of defining the phenotype of interest is not gold standard. It reasonable to speculate that at least some participants who reported seeing a psychiatrist had in fact instead seen a psychologist or counsellor. However it does not seem likely that an alternative approach to identifying subjects with moderately severe affective illness would have produced radically different results. Focussing on a more restricted phenotype, for example those exome-sequenced subjects who had required in-patient treatment for depression, would have produced an unrealistically small sample size.

The size of the sample is at the lower limit of what might be expected to yield significant findings from exome sequence data. On the other hand, since there are no biologically plausible genes with results just below conventional standards of significance there is no guarantee that a moderately larger sample would produce definitive results.

## Conclusion

The study fails to identify genes for which disturbance of function influences the probability of receiving psychiatric treatment. It seems unlikely that research into depression genetics will yield findings with substantial clinical impact until far larger samples become available.

## Data Availability

The raw data is available on application to UK Biobank. Detailed results with variant counts cannot be made available because they might be used for subject identification.

## Conflicts of interest

The author declares he has no conflict of interest.

## Acknowledgments

This research has been conducted using the UK Biobank Resource. The author wishes to acknowledge the staff supporting the High Performance Computing Cluster, Computer Science Department, University College London. This work was carried out in part using resources provided by BBSRC equipment grant BB/R01356X/1.

## Notes

### Competing Interest Statement

The authors have declared no competing interest.

### Funding Statement

No external funding.

### Author Declarations

Research Ethics Committee (REC; approval number: 11/NW/0382)

